# Association between high serum favipiravir concentrations and drug-induced liver injury

**DOI:** 10.1101/2021.05.03.21256437

**Authors:** Hitoshi Kawasuji, Yasuhiro Tsuji, Chika Ogami, Yusuke Takegoshi, Makito Kaneda, Yushi Murai, Kou Kimoto, Akitoshi Ueno, Yuki Miyajima, Yasutaka Fukui, Kentaro Nagaoka, Ippei Sakamaki, Yoshitomo Morinaga, Yoshihiro Yamamoto

**Author notes:** Correspondence: Yamamoto Yoshihiro, Department of Clinical Infectious Diseases, Graduate School of Medicine and Pharmaceutical Sciences, University of Toyama, 2630 Sugitani, Toyama, 930-0194, Japan.

## Abstract

**Objectives:** This study aimed to evaluate the incidence and pattern of favipiravir-induced liver injury and the potential association between serum concentrations in hospitalized COVID-19 patients.

**Patients and methods:** We retrospectively reviewed laboratory-confirmed patients with COVID-19 for whom serum favipiravir trough concentration (*C*_min_) was measured under steady-state conditions during treatment. All patients were administered 1800 mg of favipiravir twice daily on the first day and 800 mg twice daily from the second day.

**Results:** Thirty observed favipiravir concentrations were collected from nine patients. Of these, favipiravir-induced liver injury developed in three patients after 13 (11–14) days from the initiation of therapy, with two classified as cholestatic and one hepatocellular injury, with a score of four (possible), seven (probable), and three (possible) based on the CIMOS/RUCAM scoring system. Median (range) favipiravir *C*_min_ at steady state was found to be significantly higher in patients with liver injury at 66.4 (47.8–72.4) mg/L than in those without injury at 12.8 (9.4–21.8) mg/L (*P =* 0.028).

**Conclusions:** Higher favipiravir serum concentrations were observed in patients who developed favipiravir-induced liver injury than in those who did not. As the variations in favipiravir concentrations between patients were large, personalized optimal dosing strategies may be needed for safe use.

## 1. Introduction

Coronavirus disease (COVID-19) caused by severe acute respiratory syndrome-coronavirus 2 (SARS-CoV-2) has rapidly spread to more than 200 countries, with more than 1.4 hundred million confirmed cases and 3.0 million deaths to date [1]. Various drugs have been used to treat patients with COVID-19, and their efficacy has been evaluated to identify a breakthrough therapy.

Favipiravir is a modified pyrazine analog that inhibits RNA-dependent RNA polymerases, which ultimately prevents viral transcription and replication of RNA viruses. Favipiravir, which was initially developed by Toyama Chemical Co., Ltd., as a drug to treat the influenza virus infection, is expected to exhibit antiviral activity against SARS-CoV-2; however, information about its efficacy, particularly its related pharmacokinetics, remains limited.

The recommended dosing protocol for favipiravir in COVID-19 is 1800 mg twice a day on day 1 and 800 mg twice a day from day 2 to 10 days (maximum 14 days). Although favipiravir is regarded as safe in various studies, the risks of teratogenicity, hyperuricemia, and drug fever should be considered [2, 3]. Elevated blood uric acid levels are a frequent side effect of favipiravir. However, blood uric acid levels normalize quickly after discontinuation of favipiravir, and few symptoms due to hyperuricemia have been observed in most studies [4]. Subsequently, abnormal liver tests and/or liver injury are also important adverse effects observed frequently in some clinical trials [5, 6], which sometimes lead to discontinuation of therapy; however, case reports on favipiravir-induced liver injury are limited, and no previous study has reported an association between favipiravir-induced liver injury and serum concentrations [7].

The present study aimed to investigate the variations in favipiravir serum concentrations during treatment between individual patients and evaluate the incidence and pattern of favipiravir-induced liver injury, and the potential association between serum concentrations in laboratory-confirmed hospitalized patients with COVID-19.

## 2. Patients and methods

### 2.1 Study design

Clinical data, including favipiravir concentrations, were collected from laboratory-confirmed patients with COVID-19 who were admitted to Toyama University Hospital from April 2020 to February 2021 and treated using favipiravir. Patients with at least one favipiravir serum *C*_min_ measured under steady-state conditions, a minimum of 48 h after favipiravir initiation, during favipiravir treatment were included in this study. All patients were administered 1800 mg of favipiravir twice daily as a loading dose on day 1 and 800 mg twice daily as a maintenance dose from day 2 and thereafter.

### 2.2 Data Collection

A retrospective chart review was performed for all individuals in the study to identify basic demographic and clinical characteristics, medical history and comorbidities, clinical presentation and severity, possible adverse effects of favipiravir, treatment duration, and concomitant medications. Severity was divided into five categories: asymptomatic; mild (symptomatic patients without pneumonia); moderate (pneumonia patients without required oxygen supplementation); severe (required oxygen supplementation); and critically ill (requiring invasive mechanical ventilation, shock, and/or multiple organ dysfunction). Hematological and serum chemistry analyses performed during hospitalization, even after the end of favipiravir treatment, were retrieved and compared over time.

### 2.3 Assessment of hyperuricemia and favipiravir-induced liver injury

Regarding the definition of liver injury, the laboratory threshold criteria used to identify episodes of drug-induced liver injury and their clinical pattern (hepatocellular, cholestatic, or mixed) were based on international Drug-Induced Liver Injury (DILI) Expert Working Group Standards [8]. For hepatocellular liver injury, we required liver chemistry thresholds of alanine transferase (ALT) ≥ 5 × upper limit of normal (ULN) and R□≥□5, where the R-value was calculated as (ALT/ALT ULN)/(alkaline phosphatase [ALP]/ALP ULN). The cholestatic liver injury required ALP ≥ 2 × ULN and R□≤□2, and the mixed liver injury required ALT ≥ 5 × ULN and 2□<□R□<□5 [8]. The earliest liver chemistry elevations meeting the threshold criteria were used to determine the clinical pattern and liver injury date. The ALP values measured using the Japan Society of Clinical Chemistry method (x) were converted to the International Federation of Clinical Chemistry and Laboratory Medicine method (y) using the following formula: y=0.35x. The Roussel Uclaf Causality Assessment Method (RUCAM), which is the most widely used method to assess causality in drug-induced liver injury, was used to assess the causality between favipiravir and liver injury [9].

### 2.4 Determination of favipiravir concentration

Steady-state serum *C*_min_ was defined as the total concentration collected 9–15 h after the dose and before administration of favipiravir at ≥ 48 h after favipiravir initiation. Favipiravir concentrations were determined using residual serum samples from the clinical examination, and the serum samples collected from the patients were stored in a freezer (−80°C) until measurement. The times of oral administration and blood collection were carefully checked, and samples deemed inappropriate were excluded from the analysis. Favipiravir concentrations in the serum were determined using high-performance liquid chromatography (Shimadzu Corp., Kyoto, Japan). Favipiravir (CAS No.: 259793-96-9) was purchased as a bulk powder from Cosmo Bio Co., Ltd. (Tokyo, Japan). Separation was carried out on an aminopropyl column (Unison UK-Amino, 150 × 3 mm, 3 μm; Imtakt Co., Kyoto, Japan), and 0.1% acetic acid solution was used as the mobile phase. The lower limit of quantification was 0.5 mg/L (coefficient of variation < 10%), and the lower limit of detection was 0.1 mg/L.

### 2.5 Statistical analysis

Continuous and categorical variables are presented as median (interquartile range) and n (%), respectively. We used the Mann–Whitney U test, χ^2^ test, or Fisher’s exact test to compare the differences between patients with and without favipiravir-induced liver injury. Data were analyzed using JMP Pro version 15.0.0 software (SAS Institute Inc., Cary, NC, USA).

### 2.6 Ethics approval

This study was performed as per the Declaration of Helsinki and was approved by the Ethical Review Board of the University of Toyama (approval numbers: R2019167 and R2020146) and Nihon University (School of Pharmacy, approval number: 20-005). Written informed consent was obtained from all patients.

## 3. Results

### 3.1 Demographics and clinical characteristics

Demographics and clinical characteristics of all nine patients (seven men and two women) treated with favipiravir are summarized in Tables 1 and 2. The median age of the patients was 62 years (range, 53–90 years). Hypertension was the most common comorbidity, followed by dyslipidemia and diabetes. There were no patients with the hepatic disease with impaired liver function; however, two patients demonstrated fatty liver on computed tomography at admission. Three patients habitually ingested alcohol, which was defined as > 2 drinks per day (> 14 units/week) for women and > 3 drinks per day (> 21 units/week) for men, and 10 g ethanol for each drink. Regarding the severity of COVID-19, two patients had moderate disease, five had severe disease, and the remaining two patients were critically ill. All patients were administered 1800 mg of favipiravir twice daily as loading on day 1 and 800 mg twice daily as a maintenance dose from day 2 and thereafter. Favipiravir administration was discontinued in three patients due to favipiravir-induced liver injury in patient 3 and suspected favipiravir-induced drug fever in two patients (patients 2 and 8).

**Table 1.** Demographic and clinical characteristics of the nine patients in this study.

| Patient number | Age range (year) | Sex | Weight (kg) | Underlying comorbidities | Liver disease | Habitually ingested alcohol (drinks per day, 10 g ethanol for each drink) | Severity of COVID-19 | The type of favipiravir-induced liver injury, the occurrence days after favipiravir initiation (days) | Duration of favipiravir administration (days) | Reason for favipiravir discontinuation |
| --- | --- | --- | --- | --- | --- | --- | --- | --- | --- | --- |
| 1 | 60–69 | Male | 74.0 | Hypertension, dyslipidemia, diabetes, primary aldosteronism | - | - | Severe | - | 14 | - |
| 2 | 80–89 | Male | 68.0 | Hypertension, cerebral infarction, hyperuricemia | - | - | Severe | - | 7 | Suspected drug fever |
| 3 | 50–59 | Male | 49.5 | Hypertension, glaucoma | - | - | Critically ill | Cholestatic, 11 | 14 | - |
| 4 | 90–99 | Female | 48.3 | Hypertension, Alzheimer-type dementia, ovarian tumor | - | 2 | Severe | Cholestatic, 13 | 13 | Liver injury |
| 5 | 50–59 | Male | 78.0 | Hypertension, dyslipidemia, diabetes, hyperuricemia, sleep apnea syndrome | Fatty liver | 4 | Severe | Hepatocellular, 14 | 14 | - |
| 6 | 70–79 | Female | 62.0 | Hypertension, dyslipidemia | - | - | Critically ill | - | 14 | - |
| 7 | 70–79 | Male | 82.0 | None | Fatty liver | - | Severe | - | 14 | - |
| 8 | 50–59 | Male | 68.0 | Hypertension | - | - | Moderate | - | 7 | Suspected drug fever |
| 9 | 50–59 | Male | 69.2 | None | - | 4 | Moderate | - | 10 | - |

**Table 2.**
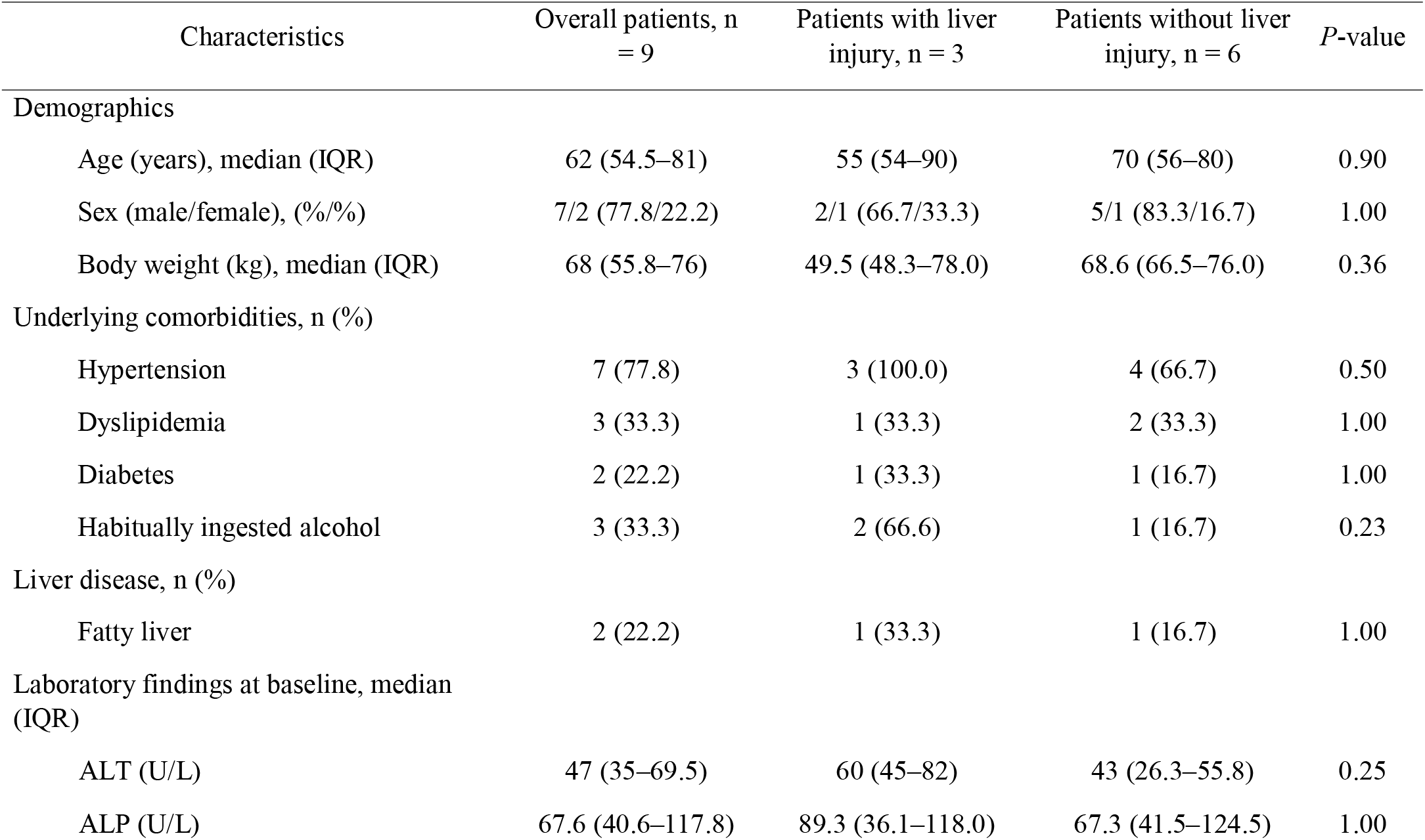
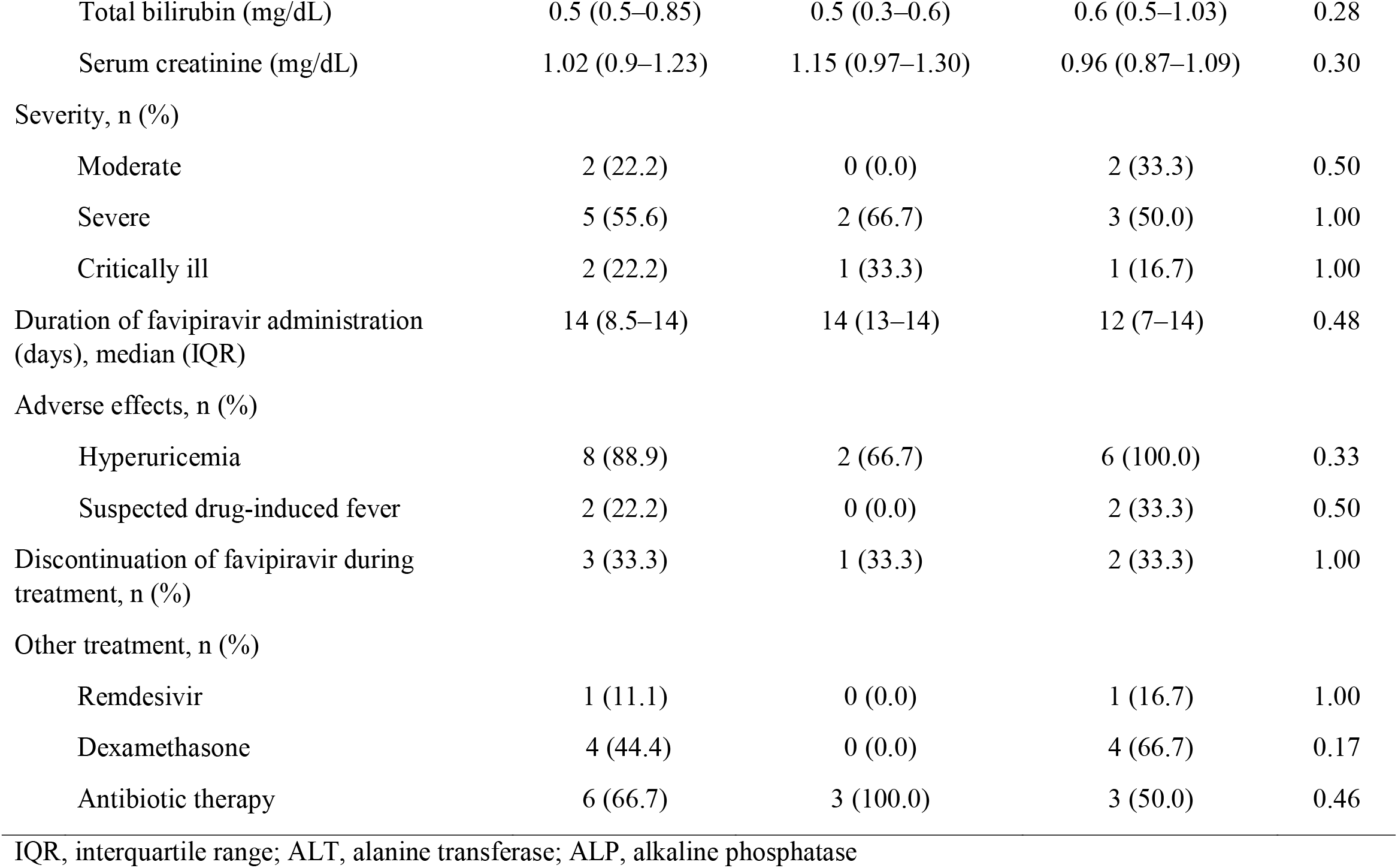
Univariable analysis of clinical characteristics between patients with or without favipiravir-induced liver injury.

Baseline laboratory findings before the administration of favipiravir are also described in Table 2. There were no significant differences between patients with and without liver injury in terms of patient demographics and clinical characteristics.

### 3.2 Favipiravir-induced liver injury and association with trough concentrations

Of the nine patients, favipiravir-induced liver injury developed in three patients during treatment. The severity of liver injury in these three episodes (patients 3, 4, and 5) assessed using the DILI severity index was mild with two classified as cholestatic (patients 3 and 4) and one as hepatocellular injury (patient 5), with a score of four (possible), seven (probable), and three (possible) based on the Research Center of Integrative Molecular Systems (CIMOS)/RUCAM scoring system. Other medications are also being considered; however, there has been no obvious relevance based on the clinical course. Among the three patients with favipiravir-induced liver injury, the median (range) time from the initiation of therapy to the development of liver injury was 13 (11–14) days. The liver injury occurred during the remission of COVID-19 in all three patients, and the RT-qPCR results using nasopharyngeal swabs obtained 1–4 days after the development of liver injury, were negative; however, were confirmed after the end of favipiravir administration. One of three patients required discontinuation of favipiravir before the planned treatment duration; however, abnormal liver blood test results gradually improved after the end of favipiravir treatment in all patients. Median (range) favipiravir *C*_min_ at steady state was found to be significantly higher in patients with liver injury at 66.4 (47.8–72.4) mg/L than in those without injury at 12.8 (9.4–21.8) mg/L (*P* = 0.028; Figure 1).

**Figure 1.**
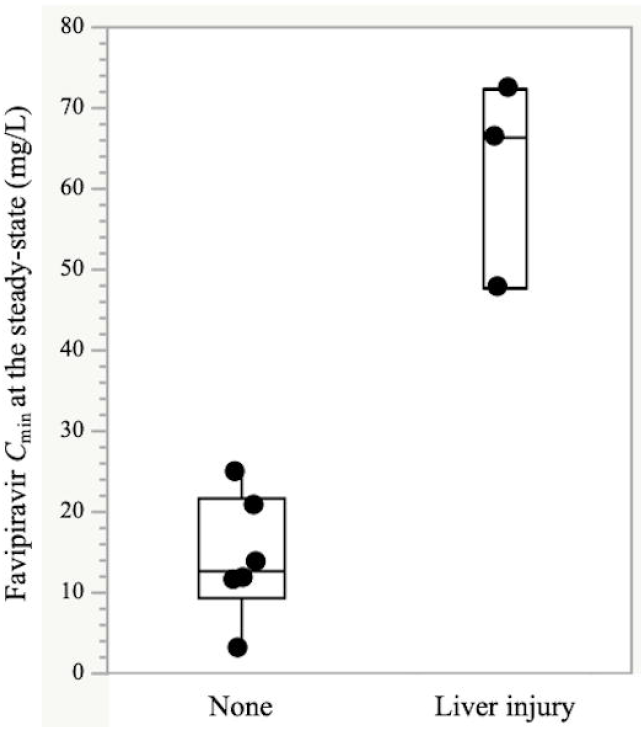
Boxplots of *C*_min_ of the patients with and without favipiravir-induced liver injury. For each boxplot, the horizontal line across the box within each box represents the median, each box represents the range between the 25th and 75th percentiles, the two whiskers represent the minimum and maximum values that are within 1.5 × interquartile range (IQR), and points beyond the whiskers represent outliers. Closed circles represent the mean *C*_min_ values at the steady-state of each patient.

### 3.3 Variations in serum favipiravir trough concentrations

A total of 30 observed favipiravir trough concentrations were collected from nine patients. The median (range) *C*_min_ was 20.8 (3.1–72.4) mg/L with an obvious inter-individual variation. In addition, the median concentrations in patient 3 decreased from 115.5 (55.5–144.8) mg/L to 5.4 (3.8–44.6) mg/L after introducing continuous hemodiafiltration (CHDF), along with the decrease of serum uric acid. Variations in favipiravir versus time are shown in Figure 2.

**Figure 2.**
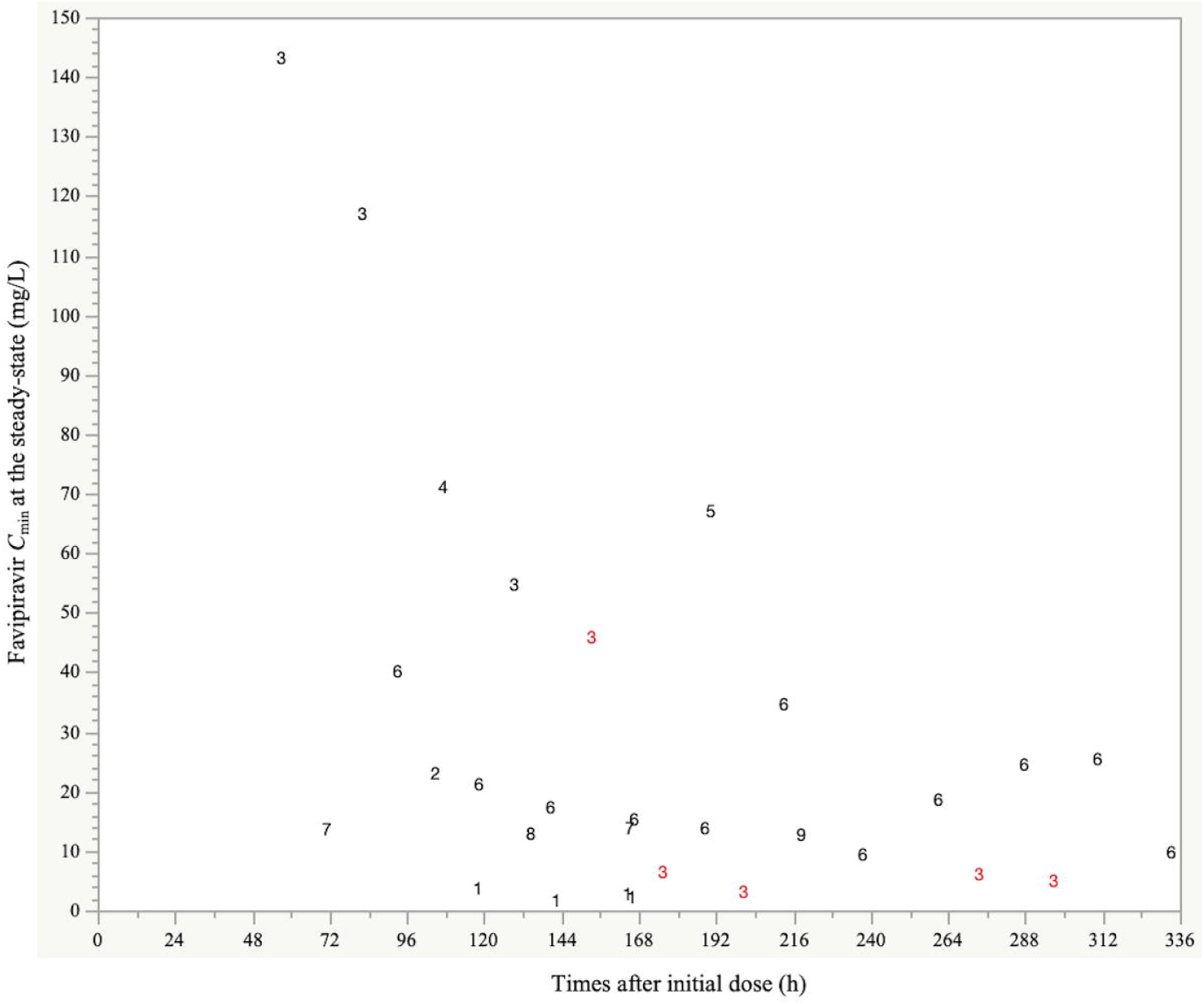
Variations of favipiravir concentrations versus time plots. Numbers represent each patient number, and the red color represents favipiravir concentrations after introducing continuous hemodiafiltration (CHDF).

## 4. Discussion

Elevated liver function tests and/or liver injury are important adverse effects of favipiravir observed frequently in clinical trials [5, 6]. Since abnormal liver function tests (LFTs) are affected by various factors, including concomitantly administered drugs, underlying liver disease, and disease severity, case reports of favipiravir-induced liver injury are still few [7] and no previous study has reported an association between favipiravir-induced liver injury and serum concentrations.

The clinical trial US109, a human study of favipiravir that looked specifically at hepatic impairment, reported that mild and moderate hepatic impairment (Child-Pugh Turcotte score A and B) resulted in 2.1-and 2.0-fold increases in favipiravir exposure, respectively [10]. It may appear that high serum concentration was not the cause of favipiravir-induced liver injury, but subsequent results of favipiravir administration in patients with hepatic impairment. However, high favipiravir serum *C*_min_ was observed before the occurrence of favipiravir-induced liver injury, there were no patients with hepatic impairment and abnormal LFTs at baseline, and elevated liver function tests were recovered after discontinuation or end of favipiravir during hospitalization.

Another factor that should be considered is the potential role of liver damage caused by SARS-CoV-2. The incidence of LFT abnormalities in patients with COVID-19 varies among studies [11-13]; however, it may be present in up to half of the hospitalized patients with SARS-CoV-2 [14, 15]. Previous studies have indicated that the hepatocellular pattern is the most frequent LFT abnormality [11-13]. These alterations have largely been reported as mild-to-moderate elevations in serum transaminases. In contrast, Yeoman et al. reported that 31% of 318 hospitalized patients with COVID-19 had one or more abnormal liver function tests and had abnormal admission in 64%, and cholestatic patterns dominated [15]. These gaps still exist, due to the largely unknown pathogenesis of liver damage by SARS-CoV-2, and other potential confounding factors, including systemic organ failure in critically ill patients and the effect of the administered drugs, also preclude detailed analysis of the causes of abnormal LFTs. The present study included five severe and two critically ill patients; however, liver injury in three patients (two severe and one critically ill) occurred not on admission, but after the clinical remission, inconsistent with the liver injury as a consequence of the systemic inflammation developed in response to the SARS-CoV-2. Other concomitant medications were also considered; however, no improvement was observed after discontinuation of the medications, and the relevance was difficult to determine.

To the best of our knowledge, there is only one case report of favipiravir-induced liver injury, despite the high frequency of abnormal LFTs and/or liver injury in clinical studies of favipiravir among patients with COVID-19 [7]. Yamazaki et al. [7] reported that a 73-year-old patient with a history of alcoholic hepatitis received favipiravir at 6000 mg/day on the first day and 2400 mg/day from the second day for 14 days; however, subsequently developed a favipiravir-induced cholestatic liver injury with a score of 6 (possible) based on the R factor and CIMOS/RUCAM scoring system [9]. This case report also suggested that favipiravir exposure may have contributed to drug-induced liver injury; however, favipiravir concentrations were not measured. In the present study, three episodes were classified as two cholestatic and one hepatocellular liver injury, with a score above three (possible).

Favipiravir may also possibly cause liver damage from the viewpoint of its chemical structure. Pyrazinamide, an antituberculosis drug, has a typical side effect of hepatotoxicity, with unclear mechanism; however, is dose-dependent [16]. Favipiravir is structurally quite similar to pyrazinamide and may be regarded as a potentially hepatotoxic drug in a dose-dependent manner.

The number of patients involved was quite small, which was a limitation of this study. Thus, representativeness is relatively insufficient, and the samples can only represent the general situation to a certain extent. However, our findings indicate a difference in serum favipiravir concentrations between patients with and without favipiravir-induced liver injury. Additionally, large variations in favipiravir concentrations were observed (steady-state *C*_min_: 3.1–72.4 mg/L) in this study of patients without renal and underlying hepatic impairment. The *C*_min_ values in patient 3 decreased from 115.5 (55.5–144.8) mg/L to 5.4 (3.8–44.6) mg/L after introducing CHDF. Although the influence of continuous renal replacement therapy (CRRT) on the clearance of favipiravir is unclear, this case suggests that evaluating the influence of CRRT could be significant for considering the optimum dose of favipiravir. These findings indicate that a pharmacokinetic multinational study should be performed with larger samples to reveal the significant covariates and elucidate the potentially exposure-dependent favipiravir-induced liver injury.

## 5. Conclusion

Higher favipiravir serum concentrations were observed in patients who developed favipiravir-induced liver injury than in those without injury. As the variations in favipiravir concentrations between patients were large, pharmacokinetic and pharmacodynamic analyses should be performed in larger populations to identify the optimum favipiravir therapy and evaluate the association between adverse effects and favipiravir serum concentrations.

## Data Availability

N/A

## Acknowledgments

We thank Rei Yasukochi for supporting the measurements of favipiravir concentrations.

## Author contributions

H.K. and Y.Y. contributed to the acquisition of data, participated in study design, analyzed and interpreted the data, and drafted the manuscript. Y.T. (Yasuhiro Tsuji) and C.O. contributed to the acquisition of data, the serum favipiravir concentration measurement, and edited the manuscript. H.K., Y.T. (Yusuke Takegoshi), M.K., Y.M. (Yushi Murai), K.K., A.U., Y.M. (Yuki Miyajima), Y.F., K.N., I.S., and Y.Y. took care of the patients. Y.M. (Yoshitomo Morinaga) perfomed SARS-CoV-2 RT-qPCR. All authors have read and approved the final manuscript.

